# A conversational agent for providing personalized PrEP support – Protocol for chatbot implementation and evaluation

**DOI:** 10.1101/2025.05.02.25326894

**Authors:** Fatima Sayed, Albert Park, Patrick S. Sullivan, Yaorong Ge

**Author notes:** **Corresponding author:** Albert Park will handle correspondence at all stages of refereeing and publication.

## Abstract

**Background:** Chatbots have the potential to reduce barriers to pre-exposure prophylaxis (PrEP), including lack of awareness, misconceptions, and stigma, by providing anonymous and continuous support. However, in the context of PrEP chatbots are still nascent; they lack personalized informational expertise, peer experiential expertise, and human-like emotional support to promote PrEP uptake and retention. Tailoring information, providing relatable peer experiences, and offering effective emotional support are all crucial for increasing engagement, influencing health decisions, and fostering resilience and well-being.

**Objective:** In this paper, we describe the iterative development of a RAG chatbot for providing personalized information, peer experiential expertise, and human-like emotional support to PrEP candidates.

**Methods:** We employed an iterative design process consisting of two phases – prototype conceptualization and iterative chatbot development. In the conceptualization phase, we identified real-world PrEP needs and designed a functional dialog flow diagram for PrEP support. Chatbot development included developing 2 components – a query preprocessor and a RAG module. The preprocessor uses the Segment Any Text (SAT) tool for query segmentation and a Gemma 2 fine-tuned support classifier to identify informational, emotional, and contextual data from real-world queries. To implement the RAG module, we used information retrieval techniques, employing Sentence-BERT (SBERT) embeddings with cosine similarity for semantic similarity and performing topic matching to identify topically relevant documents based on query topic to support document retrieval. Extensive prompt engineering is used to guide the large language model (LLM), Gemini-2.0-Flash, in generating tailored responses. We conducted 10 rounds of internal evaluations to assess the chatbot responses based on 10 criteria: clarity, accuracy, actionability, relevancy, information detail, tailored information, comprehensiveness, language suitability, tone, and empathy. The iterative feedback was used to refine the LLM prompts to enhance the quality of chatbot responses.

**Results:** We developed a RAG chatbot and iteratively refined it based on the internal evaluation feedback. Prompt engineering is essential in guiding the LLM to generate responses tailored to information, experiential, and emotional user needs. We found that prompt effectiveness varied with task complexity; this was likely due to LLM sensitivity to the structure of prompts and to linguistic variability. For tasks requiring diverse perspectives, fine-tuning LLMs on annotated datasets provided better results compared to few-shot prompting techniques. Prompt decomposition and segmenting prompt instructions helped improve comprehensiveness and relevancy for complex and long queries. For tasks with high decision variance, condensed prompts that summarize the main concept or idea were more effective in reducing ambiguity in LLM decisions compared to decomposed prompts.

**Conclusion:** Our RAG chatbot leverages social media data to provide personalized information, peer experiences, and human-like emotional support; these elements are essential in effectively reducing PrEP misconceptions and promoting self-efficacy. Further analysis incorporating expert and user feedback will be conducted to help validate and improve the chatbot’s potential.

## Introduction

Increasing the uptake of pre-exposure prophylaxis (PrEP) [1–3] is a key strategy in the Ending the HIV Epidemic (EHE) initiative [4]. However, social and structural barriers to HIV treatment seeking behaviors continue to impact health outcomes and drive inequities [5–7]. PrEP adoption has been hindered in the United States by lack of PrEP knowledge and prevailing anticipated and experienced stigma and discrimination [5,7–9]. Personal experiences shared by peers can improve the awareness [10,11], self-efficacy and intrinsic motivation [12] of people considering PrEP use, mitigating their stigmatized beliefs [10] and informing health decisions [10,13]. People without financial means or ready access to providers experience limited access to healthcare [14,15]. Service deserts result in a lack of physically proximate service providers [6]; scarcity of services also disproportionately impacts those without financial needs or access [16]. Equitable PrEP uptake requires facilitating access to PrEP resources while meeting the diverse needs of PrEP candidates [16]. Conversational agents (i.e., chatbots) have the potential to provide anonymous round-the-clock assistance in finding a PrEP provider, answering questions about PrEP and providing emotional support. These services can help to overcome geographical and access barriers to seeking PrEP care.

### HIV and PrEP Chatbots

Chatbots have demonstrated potential in disseminating HIV information [17,18], self-testing [17,19,20], access to and uptake of HIV prevention and care [21–23], and making positive behavioral changes relevant to HIV prevention and care (e.g., self-disclosure of HIV status [24,25], self-management of PrEP adherence [17,21,26]). AI technologies can be useful in developing and operating chatbot services, but their contribution is variable depending on the risk and complexity associated with health infrastructure. This is because risk and complexity influence the adoption of chatbots by users for improving health conditions [27]. Rule-based chatbots, which use scripted text using simple rules (i.e., keyword identification and pattern matching techniques) are most commonly used to provide information [18,28] and support for self-management of health behaviors (e.g., setting up reminders) [21,24]. More advanced chatbots use information retrieval systems [22] and knowledge graphs [20] to help users identify HIV risk and navigate relevant information to mitigate barriers to PrEP uptake. For more complex needs such as identifying disorders and providing behavioral support (e.g., for PrEP retention), chatbots use hybrid techniques that employ AI tools (e.g., machine learning [19,20,22] and natural language processing [20]) to learn patterns from previous conversations [29]. These approaches can also detect emotions [29] and perceive user characteristics based on past interactions [20], and can integrate dialog rules for response generation. Two protocols for studies mention the use of a hybrid LLM (HumanX) chatbot for promoting PrEP uptake and utilization [23,26] and for providing HIV related mental health support. [21]. However these studies do not specify details of the chatbot implementation. Such rule-based and hybrid healthcare chatbots are often criticized for their limited flexibility [30,31] and diversity of content [30–35], and insufficient contextual understanding [36,37] and personalization [22,38]. Such chatbots have also been reported to lack human-like emotional support [39], and this limits long-term user engagement and chatbot effectiveness [40].

To date, no study details the implementation of a large language model (LLM)-based chatbot for providing various HIV and PrEP support, including emotional, informational, and peer experiential support to facilitate access to PrEP information and promote PrEP uptake and adherence. Although LLMs can offer improved personalization, and human-like conversations [41,42], their performance can also be limited by their scope of knowledge, diversity of available data and the potential for LLM hallucinations [43,44]. LLMs are probabilistic models and can hallucinate, generating incorrect information that seem to be plausible. Prompt engineering [45] and retrieval augmented generation (RAG) [44] techniques improve LLMs’ performance in terms of accuracy [46,47], relevancy [46,48] and user personalization [49,50], which can enhance user trust in chatbot provided support [51–53]. Using prompt engineering, LLMs can be guided to improve the precision and accuracy of generated responses [52]. RAG chatbots have indicated improved accuracy (34%) [45] and reduced hallucination (26.5%) [44] in generated responses compared to LLM question-answering bots. However, the development of chatbots based on LLM and RAG techniques is still nascent in the context of HIV and PrEP support and has not been implemented to address implicit user needs [44], answer long contextual queries [44] and generate personalized responses [54]. Addressing these issues in RAG chatbots can lead to providing accurate, flexible, personalized and human-like responses to promote PrEP uptake.

Available literature [56,57,111] and our prior work have identified human-like emotional support, components as essential elements to support behaviors change for HIV medication taking and PrEP uptake. However, no prior work has developed RAG chatbots to provide personalized information and peer experiential expertise (e.g., user stories) [55]. Existing HIV and PrEP chatbots focus on providing informational support [18,21,28] and emotional support according to therapeutic guidelines [26]. Effective support provided by humans relies on emotional support (i.e., human-like emotional support) [58,59] and peer experiential expertise (i.e., peer experiential support) [55,60], elements which are absent in existing healthcare chatbots. Providing effective emotional support first requires understanding and communicating complex human emotional experiences [61–63]. The Willcox’s Feeling Wheel facilitates this process by providing a related vocabulary for 6 emotion categories [63]. To better reflect the real-world user queries that often seek practical user experiences (peer experiential expertise) [64], PrEP chatbots should provide peer experiential expertise support, which might enhance self-coping skills and emotional confidence through pragmatic narratives of others in similar situations, leading to improved quality of life [55,60]. Additionally, chatbots need to support back-and-forth dialog exchanges, similar to real-world help seeking behaviors, to facilitate clarification and understanding of complex concerns [65].

### Protocol Goals

This protocol describes the development of a RAG chatbot in two phases. Phase 1 describes conceptualizing the chatbot architecture, and Phase 2 includes iterative development of the RAG chatbot to provide personalized information, peer experiential expertise, and human-like emotional support. Before prototype development, we conducted a separate study analyzing PrEP-related online social support exchanges to identify the social needs of PrEP candidates to conceptualize the chatbot architecture. Results of this study will be reported in a separate manuscript. We demonstrate the iterative development process focusing on prompt engineering, which is an integral part of the LLM RAG chatbot implementation. We also discuss the implications and potential of our chatbot in improving PrEP uptake. The heuristic evaluation study is exempt from IRB review because evaluators provide their opinions and feedback solely on the quality of chatbot responses and, as such, are not considered study participants.

## Methods

### Phase I: Prototype Conceptualization

To inform goals of our RAG chatbot prototype, we analyzed 3020 publicly available Reddit conversations focused on PrEP from April 2011 to March 2024. We focused on the informational (including peer experiential expertise), and emotional (i.e., psychosocial) support because these are the two main types of support that are exchanged in this forum and because these have the potential to impact broader behavioral trends related to PrEP use [66]. Our database for RAG aligns with the previous literature [106,107]. We found that over 80% of user PrEP needs included informational and emotional need items. Informational needs centered on PrEP facts and related experiences, and emotional needs focused on reassurance, HIV/PrEP concerns sharing, empathy, and sympathy, based on the psychosocial constructs of Social Support Behavior Codes (SSBC) [67,68]. PrEP candidates’ emotions can be described with Willcox’s Feeling wheel [63], including Sad, Mad, Scared, Joyful, Peaceful, and Powerful.

Real-world user queries were multifaceted consisting of multiple informational and emotional needs. Chatbots must (1) pre-process each input query into semantic segments; (2) generate responses for each segment and (3) combine them into a final response. Based on literature [106,107] and our prior work, our chatbot was designed to: (a) generate personalized responses; (b) provide accurate and comprehensive information to user queries; (c) provide peer experiential expertise support; and (d) respond empathetically to the emotions expressed by the user. Functional dialog flow diagrams were created (e.g., Figure 1) to visually communicate with our team members and developers how the chatbot would achieve these aims.

**Figure 1.**
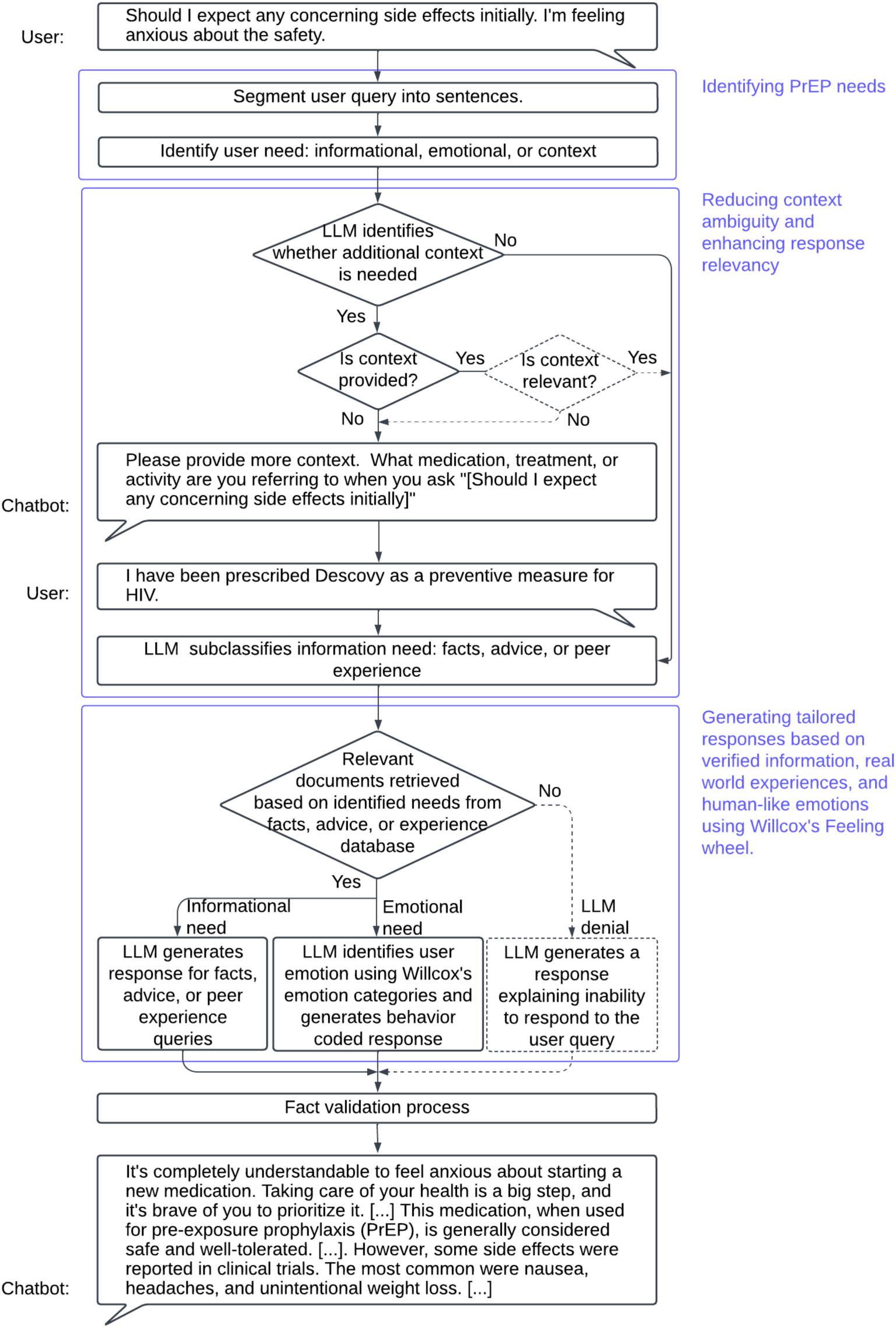
An example of a generated response using a paraphrased real-world query from Reddit where a user seeks factual informational and emotional support. The input query is ambiguous, and lacking sufficient context thus, to handle the ambiguous query, the chatbot requests additional context. The dotted line in the diagram indicates ongoing updates.

To identify user needs, we created a PrEP-related dataset. This dataset was annotated with three categories: informational (including facts and peer experiential expertise), emotional, and context. Subsequently, we built a classifier to parse user inputs and categorize them into the three categories. The classifier further classified informational needs into facts and peer experiential expertise. For informational needs, we specifically designed the chatbot to provide information (e.g., facts and actionable advice) and peer experiential expertise support (i.e., other people’s experiences and opinions). For emotional needs, we designed the chatbot to identify user emotions based on Willcox’s Feeling Wheel [63] and to generate a suitable response using relevant SSBC psychosocial constructs known to impact PrEP uptake (e.g., i.e., reassurance, understanding, empathy, sympathy, validation, relief of blame, and compliment). [56,69].

### Phase II: Iterative Prototype Development

The iterative development cycle (Figure 2) consists of four processes: 1. system architecture design - drafting initial prompts for guiding LLM response generation, 2. internal evaluation of chatbot responses, 3. modification of RAG chatbot based on internal evaluation feedback, and 4. formal evaluation by HIV experts and users.

**Figure 2.**
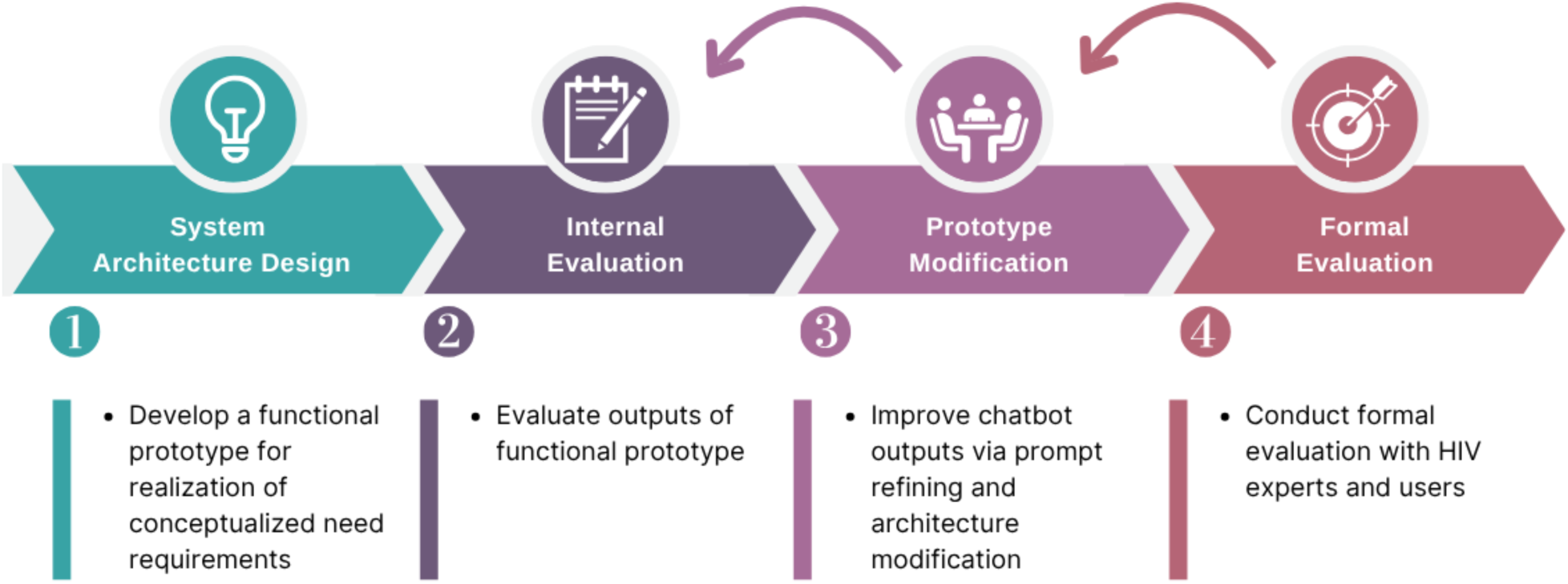
Research process of developing and refining a RAG chatbot for providing support to PrEP candidates. The arrows looping back to processes 2 and 3 indicate a recursive process in which the RAG chatbot is refined based on internal evaluation and formal evaluation.

#### System Architecture

The RAG chatbot was developed using Python programming and accessing Google Gemini API. The chatbot consists of 2 main modules (Figure 3): a preprocessor and a RAG module. The RAG, in turn, leverages two databases: a facts database and an experience database. The facts database contains verified information from factsheets, brochures, and websites on HIV and PrEP from public health agencies and private health organizations. The experience database contains Reddit user experiences and emotions related to PrEP concerns. The preprocessor extracts subqueries and context information from user input. The information retriever uses the extracted information to handle context management and information subclassification to enhance semantic retrieval, and outputs relevant documents. The LLM generator produces a personalized response using the retrieved documents, context information, and subquery.

**Figure 3.**
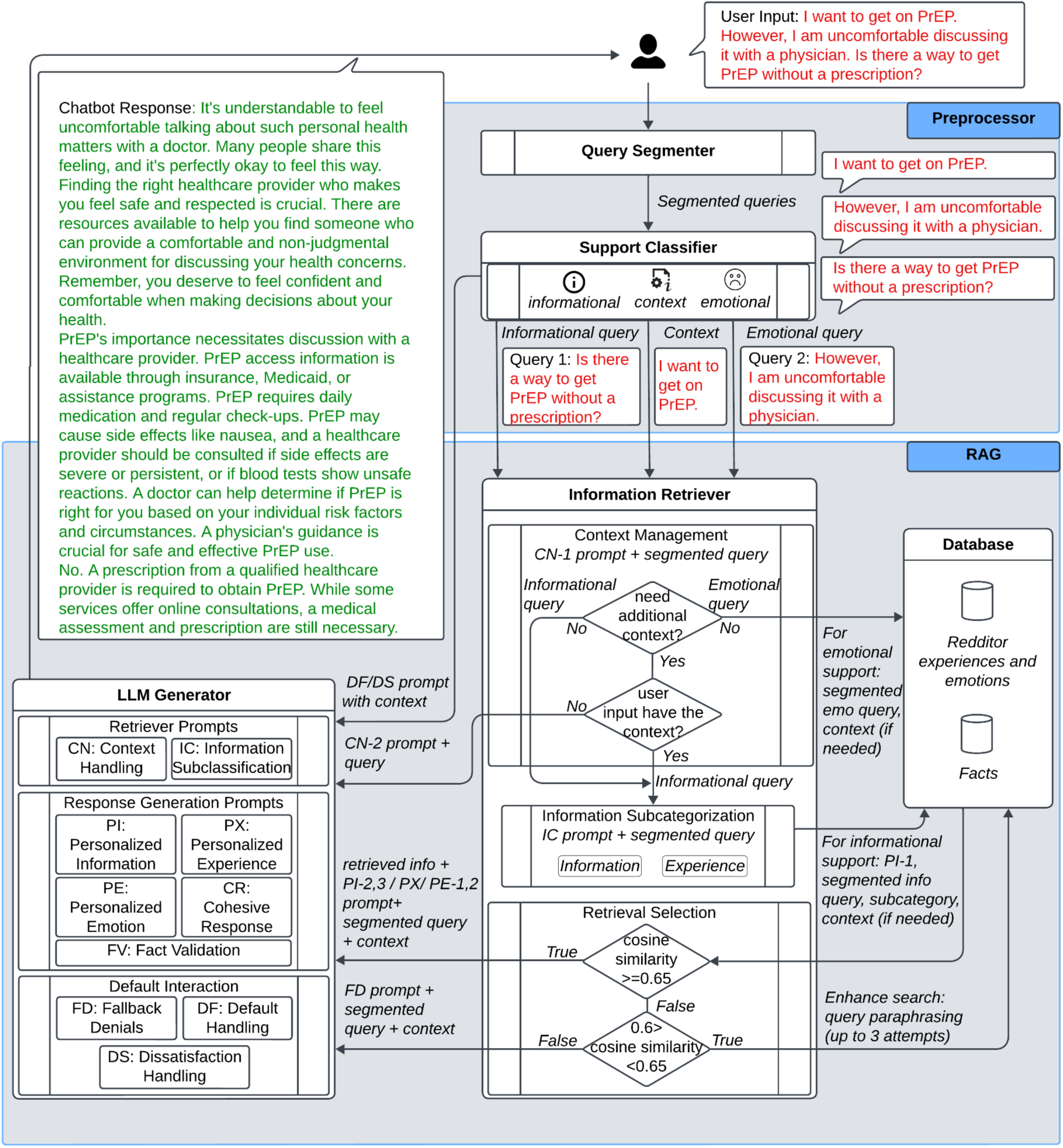
Architecture of the RAG chatbot. The preprocessor extracts user needs and context from user input. Using this information, the RAG module retrieves relevant information from the facts and experience databases to generate a response, which is then validated using the retrieved relevant information. Personalized prompts guide the LLM to generate personalized information, peer experiential expertise and human-like emotional responses for providing tailored responses.

### Preprocessor Module

The preprocessor module consists of two components: a query segmenter, and a support classifier.

#### Preprocessor: Query Segmenter

The query segmenter semantically segments a user input into sentences using the Segment-Any-Text (SAT) tool [70]. The tool is highly robust because it is minimally reliant on punctuation and space characters. This makes it an optimal choice for handling diverse writing styles and complex real-world user queries. Each segment is then passed to the support classifier.

#### Preprocessor: Support Classifier

The 3-class classifier was built by fine-tuning the Gemma-2-2B-it model on a class balanced subset (N=10,041) of the PrEP dataset. The classifier was used to classify each segmented query into one of the three categories: information, emotion, or context. Gemma is an open-source generative AI model from Google, capable of developing applications for a variety of tasks. Gemma model series are lightweight, and hardware flexible, offering low latency and balancing performance and efficiency, making it suitable for chatbot deployments [71]. We used Parameter Efficient Fine Tuning (PEFT) with standard Lora configurations (alpha=32 and rank=64) [72] on a sequential classification task for fine-tuning the Gemma models. Based on previous literature on LLM fine-tuning performance, we fine-tuned Gemma-2B-it model for 5 epochs with an early stopping patience of 2 epochs [73,74].

To select the model, we compared the performance of 6 classifiers. Two models were the Gemma-2B-it and Gemma-2-2NB models described above; the other was the Bidirectional Encoder Representations from Transformers (BERT) model. BERT is the state-of-the-art model used in text classification tasks. We trained each of the three models on two batches of data: the entire (N=32,081) PrEP-related dataset, and a class balanced subset (N=10,041) of the PrEP dataset. BERT-base-uncased model is finetuned for 5 epochs. As reported in Table 1 and Table 2., our fine-tuned Gemma 2 2B-it model demonstrated a strong performance in classifying PrEP support needs when fine-tuned on the full dataset (F1=0.86) and class balanced subset (F1=0.89). Specifically, we found that fine-tuning the Gemma 2 2B-it model on a class balanced subset indicated strong accuracy and precision in classifying emotional needs, while maintaining comparable performance in classifying informational and context queries (Table 1). In the context of classifying informational and emotional seeking support, our class unbalanced BERT model exhibits superior performance (F1-information = 0.92, F1-emotion = 0.71) compared to a previous social support seeking dataset CHQ-SocioEmo (F1-information = 0.58, F1-emotion = 0.62). This classification of user queries allows our chatbot to identify user needs, extract context, and generate tailored and emotionally relevant responses.

**Table 1.**
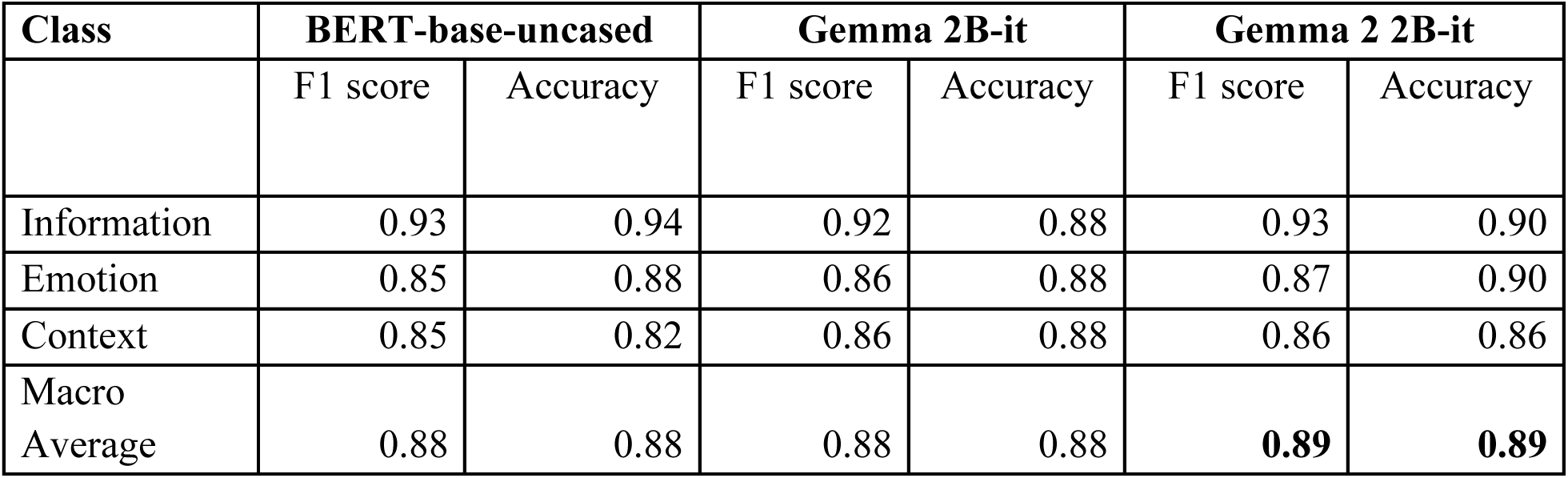
Results of an assessment of the performance of three classifiers, fine-tuned on the class balanced PrEP dataset. These classifiers are used for classifying user input into three classes. Bold values represent the best performing model.

**Table 2.** Results of an assessment of the performance of three classifiers fine-tuned on the full PrEP dataset. These classifiers are used for classifying user inputs into three classes. Bold values represent the model with best performance and value denoted as ☨ represents poor performance compared to class-balanced fine-tuned model predictions.

| Class | BERT-base-uncased |  | Gemma 2B-it |  | Gemma 2 2B-it |  |
| --- | --- | --- | --- | --- | --- | --- |
|  | F1 score | Accuracy | F1 score | Accuracy | F1 score | Accuracy |
| Information | 0.92 | 0.929 | 0.92 | 0.927 | 0.92 | 0.92 |
| Emotion<sup>†</sup> | 0.71 | 0.693 | 0.69 | 0.624 | 0.71 | 0.663 |
| Context | 0.95 | 0.948 | 0.95 | 0.964 | 0.96 | 0.966 |
| Weighted average | 0.92 | 0.92 | 0.92 | 0.922 | 0.92 | **0.93** |

### RAG Module

The RAG module consists of an information retriever and an LLM generator.

#### RAG: Information Retriever

The information retrieval process includes context management, information sub-categorization, and retrieval selection.

##### Information Retriever - Context Management

The context handler instructs the LLM (Gemini-2.0-Flash) using the context handling prompt 1 (CN-1) to determine whether queries require additional context. If no additional context is needed, only the user query was used for retrieving relevant documents. If additional context is needed, our chatbot will ask users for additional context using the context handling prompt 2 (CN-2). With additional context provided by the user, the chatbot then retrieved relevant documents using both the context and user query.

##### Information Retriever - Information Subcategorization

To generate personalized responses, the information needs were further sub-categorized (1) seeking facts or (2) request for peer experiential expertise, using the information subclassification few-shot prompt (IC) (Table 3). The identified information subcategory (i.e., facts or peer experiential expertise) was used to guide the retrieval process. We included the ‘advice’ subclassification in the few-shot examples as ongoing updates include integrating the “MedHelp” database, an online health discussion board containing expert advice related to PrEP.

**Table 3.** Prompts used for providing personalized informational, peer experiential expertise, and human-like emotional support.

| Category | Description | Prompt |
| --- | --- | --- |
| **Context Handling (CN) Prompts** | CN-1: Prompt used for identifying whether additional context is needed | [{query}]<br>Identify whether the query enclosed in square brackets above is self-contained. Provide corresponding one-word response as 'Yes' if self-contained and 'No' if not self-contained. |
|  | CN-2: Prompt used for instructing LLM to ask user for additional context | Ask user to provide more information or context in relation to the query enclosed in square brackets.<br>[{query}] |
| **Information Subclassification (IC) Prompt** | IC: Prompt for subclassifying information queries as seeking information or peer experiential expertise support | Classify the following text as seeking advice, experience, or information:<br>Text: should i take this medicine.<br>Classification: Advice<br>Text: Looking for support, personal stories, or experiences.<br>Classification: Experience<br>Text: what is PrEP<br>Classification: Information<br>Text: 'I don't know what to do.<br>Classification: Advice<br>Text: How are other people coping with PrEP side effects?<br>Classification: Experience |
|  |  | Text: Is it worth to buy Truvada?.<br>Classification: Information<br>Text: |
| **Personalized**<br>**Information (PI)**<br>**Prompts** | PI-1: Prompt used for identifying topic discussed in the query for retrieving similar documents based on topic match. | [{context} {query}]<br>Topic list = {topic_list}<br>From the above topic list, assign a single main topic that is most suitable to the text enclosed in square brackets. If none of the topics is most suitable, review the above topic list and pick a closely related topic. Provide only the topic. |
|  | PI-2: Prompt used for generating responses to informational query using the retrieved documents. | [{messages}]<br>Segment the following query and provide a detailed response to all segments in paragraphs by using **\*\*only** the information enclosed in square brackets above. Provide a single response combining all segment responses in plain paragraph format and **\*\*do not specify** the segments. Also, **\*\*Remember not to use unnecessary special characters in the response.** - {context} {query}<br>**\*\*Important instruction: Generate a response without containing sentences with phrases like 'the provided text', 'the passage', 'the document', 'the information', 'the text', 'provided information', or 'provided text'.** |
|  | PI-3: Prompt used to refine generated response. | [Query: {context} {query}<br>Response: {response text}]<br>For the user query and response pair enclosed in square brackets, check if the response is consistent with the query in details such as location, age, and gender. If the response is not consistent, paraphrase the response to ensure that the response is general and does not include any details related to location, age, gender. **\*\*Only provide the paraphrased response. If the response is consistent provide **\*\*only** the response text without the prefix Response: and without the square brackets.** |
| **Personalized**<br>**Experience (PX)**<br>**Prompt** | Prompt used for generating response to patient expertise query using the retrieved documents. | Using **\*\*only** the information enclosed in square brackets above, provide a response to the following query - {context} {query} Present the response as a third person narrative opinion, starting with something as "There are people with similar experience ....". You can also give direct speech example of the user experience. **\*\*Remember to end the response stating that the information in the response is based on other people's experience.**<br>**\*\*Important instruction: Generate a response without** |
|  |  | containing sentences with phrases like 'the provided text', 'the passage', 'the document', 'the information', 'the text', 'provided information', or 'provided text'. |
| ***Personalized Emotional (PE) Prompts*** | PE-1: Prompt used for generating response to emotional query using the retrieved documents. | Consider the following Feeling = Emotion categories:<br>Sad = Tired, bored, lonely, depressed, ashamed, guilty, sleepy, apathetic, isolated, inferior, stupid, remorseful<br>Mad = Hurt, hostility, angry, selfish, hateful, critical, distant, sarcastic, frustrated, jealous, irritated, skeptical<br>Scared = Confused, rejected, helpless, submissive, insecure, anxious, bewildered, discouraged, insignificant, inadequate, embarrassed, overwhelmed<br>Joyful = Excited, sensuous, energetic, cheerful, creative, hopeful, daring, fascinating, stimulating, amused, playful, optimistic<br>Powerful = Aware, proud, respected, appreciated, important, faithful, surprised, successful, worthwhile, valuable, discerning, confident<br>Peaceful = Nurturing, trusting, loving, intimate, thoughtful, content, thankful, secure, serene, responsive, pensive, relaxed<br>Identify the feeling expressed in the following expression based on the feelings=emotions listed above - {query}<br>For the identified feeling, generate a suitable response providing understanding, empathy, sympathy, reassurance, validation, relief of blame, or compliment, for the following user query - {context} {query}. You can also use any emotional context from the information enclosed in square brackets above to support your response. Do not use facts from the information enclosed in square brackets above to support your response. Only provide the final response without the identified feeling. Also, **Do not use unnecessary special characters in the final response. |
|  | PE-2: Prompt used for generating responses to emotional query when no relevant documents are retrieved. | Consider the following Feeling = Emotion categories:<br>Sad = Tired, bored, lonely, depressed, ashamed, guilty, sleepy, apathetic, isolated, inferior, stupid, remorseful<br>Mad = Hurt, hostility, angry, selfish, hateful, critical, distant, sarcastic, frustrated, jealous, irritated, skeptical<br>Scared = Confused, rejected, helpless, submissive, insecure, anxious, bewildered, discouraged, insignificant, inadequate, embarrassed, overwhelmed<br>Joyful = Excited, sensuous, energetic, cheerful, creative, hopeful, daring, fascinating, stimulating, amused, playful, optimistic<br>Powerful = Aware, proud, respected, appreciated, important, faithful, surprised, successful, worthwhile, valuable, discerning, confident<br>Peaceful = Nurturing, trusting, loving, intimate, thoughtful, content, thankful, secure, serene, responsive, pensive, relaxed<br>Identify the feeling expressed in the following expression based on the feelings=emotions listed above - {query}<br>For the identified feeling, generate a suitable response providing understanding, empathy, sympathy, reassurance, validation, relief of blame, or compliment, for the following user query - {context} {query}. Only provide the final response without the identified feeling. Also, **\*\*Do not use unnecessary special characters in the final response.** |
| **Fact Validation (FV) Prompt** | FV-1: Prompt used for verifying fact consistency. | Review the following list of response and document sentence pairs and identify responses that are inconsistent with the corresponding document sentences. Only provide the inconsistent response sentences with their key and value as a list of json dictionary. If all response-doc pairs are consistent, say 'All consistent'. Consistency means that all information provided in the response text is fully supported by the document text. If the response text contradicts, differs or provides partially incorrect information with respect to the document text, it is inconsistent. Start response as Response = {fact list} |
|  | FV-2: Prompt used for extracting supporting facts for inaccurate response-document pairs. | Text: [{text}]<br>For each of the response sentences in the json list below, extract corresponding sentences from the text enclosed in square brackets above, that fully support the specific response sentence. If no supportive sentence is found, replace 'document sentence' value as "" else replace the |
|  |  | 'document_sentence' value with the extracted text in the json dictionary. Only provide the json response starting as 'json []' without additional text.<br>Return **\*\*only\*\*** a valid JSON response:<br>````json { "id": "0", "response_sentence": "...", "document_sentence": "...", } {answer}` |
| ***Cohesive Response (CR) Prompt*** | Prompt used for combining responses generated for sub-queries. | [ {response text} ]<br>Smoothen the flow of information in the text enclosed in square brackets. Only provide the final response. |

##### Information Retriever - Retrieval Selection

The retriever uses Sentence-BERT (SBERT) embeddings [75] to retrieve relevant documents from the database. We used SBERT embeddings due to its bi-directional contextual understanding [75], computational efficiency [75] and superior performance in identifying semantically similar sentences, coupled with synonyms and negation lexical variations [76], which are better suited to cover various user writing styles. The retriever uses cosine similarity and LLM identifies topics for information retrieval; this was done to provide facts in informational or emotional support. We used topic matching to enhance the retrieval of a wide variety of relevant documents from the facts database, to include context rich queries that challenge semantic retrieval [77] in short query databases. We manually reviewed the facts database to identify 46 broad HIV-PrEP related topics (Textbox 1). To annotate the facts database, we instructed the LLM (Gemini-1.5-flash) model to assign the most suitable topic to each query from this list. If no relevant topic is identified, we instructed LLM to generate a closely relevant topic to have the ability to adapt to unforeseen user needs. We then manually reviewed the annotated topics to include any LLM generated new topics into the topic list and to merge semantic duplicates. We used Gemini-1.5-flash, the latest model at that time, and retained the topic lists because we manually verified all the LLM generated topics and were satisfied with the results.

**Textbox 1.** List of topics used in RAG topic document retrieval for factual support. The HIV-PrEP topic list was created by manually reviewing the facts database and updated using LLM topic assignment. Topic document retrieval from facts database was used to improve sensitivity in unstructured queries.

|  |  |  |
| --- | --- | --- |
| ‘PrEP or Pre-Exposure Prophylaxis’ | ‘Sexually Transmitted Infections (STIs)’ | ‘PEP eligibility’ |
| ‘PrEP eligibility’ | ‘Viral load or Undetectable/Untransmittable’ | ‘PrEP access and cost in UK’ |
| ‘PrEP effectiveness’ | ‘HIV treatment’ | ‘Medication adherence’ |
| ‘PrEP access and cost in Australia’ | ‘PrEP medications’ | ‘Truvada side effects’ |
| ‘PrEP access and cost’ | ‘Emtricitabine/tenofovir disoproxil fumarate’ | ‘Healthy lifestyle’ |
| ‘HIV testing’ | ‘Emtricitabine/tenofovir disoproxil fumarate administration’ | ‘Descovy’ |
| ‘HIV self test’ | ‘Emtricitabine/tenofovir disoproxil fumarate side effects’ | ‘PrEP safety and precautions’ |
| ‘PEP or Post exposure prophylaxis’ | ‘Drug interactions’ | ‘HIV Prevention Strategies’ |
| ‘PrEP administration and dosage’ | ‘Emtricitabine/tenofovir disoproxil fumarate eligibility’ | ‘Truvada’ |
| ‘Apretude or Injectable PrEP’ | ‘Anti-retroviral/HIV drug side effects’ | ‘PrEP eligibility in UK’ |
| ‘PrEP side effects’ | ‘Test disclosure and anonymity’ | ‘Descovy side effects’ |
| ‘HIV and AIDS’ | ‘Stigma and its impact on people living with HIV’ | ‘HIV/AIDS drugs’ |
| ‘Risk and transmission’ | ‘PEP access and cost’ | ‘Truvada access and cost’ |
| ‘HIV prevention strategies’ | ‘PEP side effects’ | ‘Descovy access and cost’ |
| ‘Safety and precautions’ | ‘Apretude or Injectable PrEP side effects’ | ‘Emtricitabine/tenofovir disoproxil fumarate safety and precaution’ |
| ‘Cabotegravir’ | ‘Cabotegravir side effects’ |  |

###### Retrieval for factual support

For factual support, we retrieved the top 5 documents from the facts database with a cosine similarity score >=0.65. The cosine similarity thresholds were determined based on experiments and we set k = 5 for document retrieval based on previous evaluation benchmarks [78,79]. In addition, we selected 5 topically relevant documents, using the topic identified from the 46 HIV-PrEP topics (Textbox 1). To identify the topic of an informational query, we instructed the LLM (Gemini-2.0-flash) using the HIV-PrEP topic list and the personalized information prompt-1 (PI-1) to assign one suitable topic to the user query. Retrieving documents using cosine similarity was based on the context need. If context is needed, we retrieved 3 sets of top 5 relevant documents (Algorithm 1) by comparing cosine similarity between a). the context-added query and database questions (m1), b). the context-added query and database answers (m2), and c). the context-free query and database questions (m3). Both database questions and then answers were semantically searched to improve the retrieval precision. Cosine similarity between the context-free query and database answers did not improve the retrieval precision and was therefore not used in processing the context-free queries. As the majority of the facts database consists of short questions from PrEP factsheets and brochures, the retriever often failed in retrieving relevant information for queries with additional context. If no context is needed, we retrieved the top 5 documents by only comparing cosine similarity between the context-free query and the database questions.

###### Algorithm 1.

Retrieval algorithm pseudocode for improving retrieval precision in factual queries using cosine similarity. In factual queries that require additional context, the algorithm retrieves 3 lists of documents: m1, m2, and m3. Documents m1 are retrieved by comparing cosine similarity between context-added query and database questions; documents m2 are retrieved by comparing cosine similarity between context-added query and database answers; and documents m3 are retrieved by comparing cosine similarity between context-free query and database questions. With no additional context required, the algorithm only compares cosine similarity between context-free query and database questions.

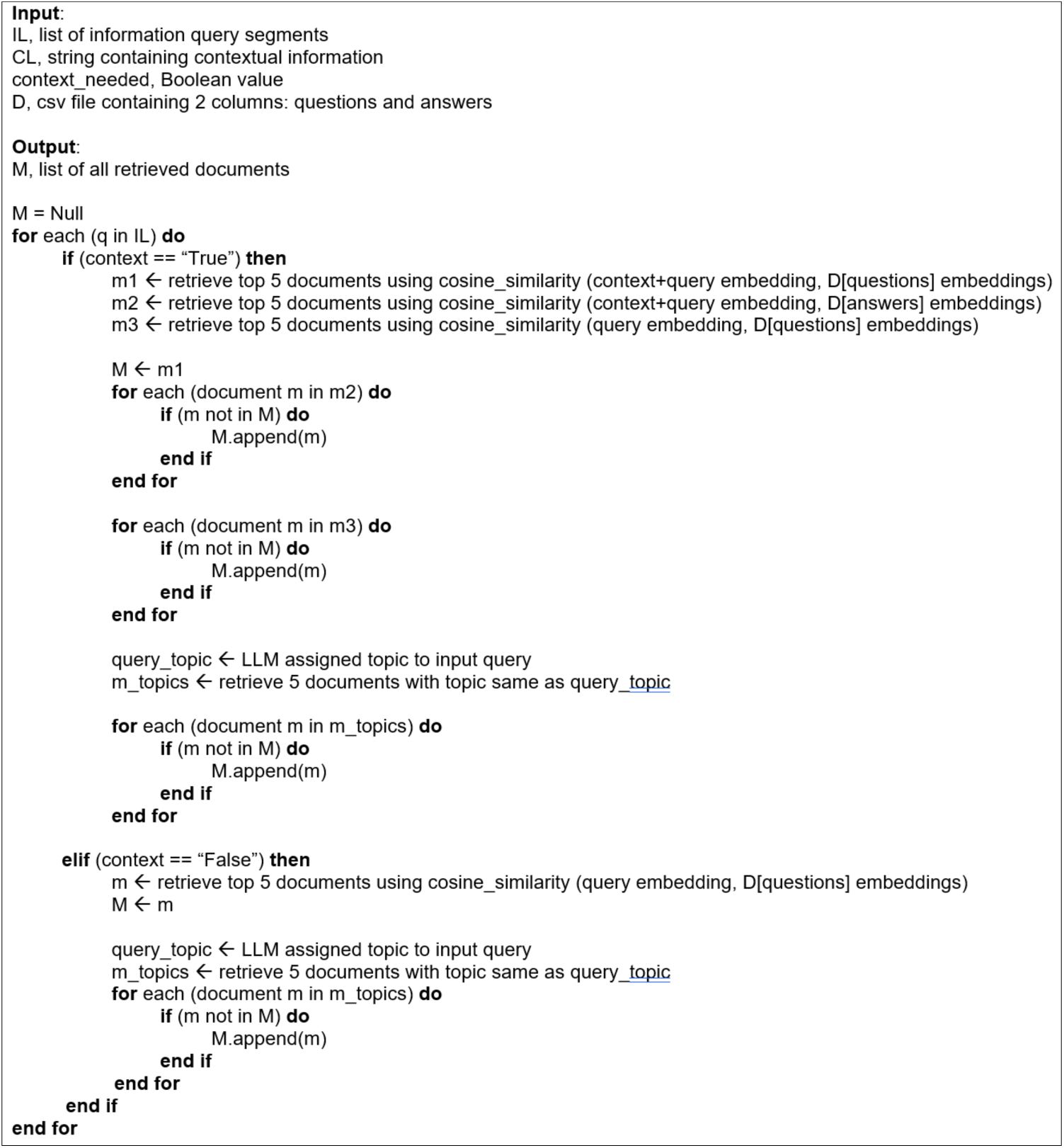

###### Retrieval for peer experiential expertise and emotional support

For peer experiential expertise support, we retrieved the top 5 documents from the experience database using cosine similarity with a score of at least 0.65. When context is needed, we separately used both the query and context for retrieval to overcome context-dependent retrieval failure. For emotional queries, the top 5 similar documents were retrieved from the experience database in addition to providing relevant facts. Similar to document retrieval for peer experiential expertise support, we retrieved the top 5 documents from the experience database using cosine similarity with at least 0.65 score and when context is needed, used both the query and context for retrieval. As information can be helpful in addressing certain HIV-PrEP emotional concerns [80], we retrieved facts relevant to the emotional query.

To improve the sensitivity, we also used up to 3 paraphrase queries with a lower cosine similarity score, between 0.6 and 0.65. For paraphrasing, we used the T5 paraphrase generator from the Transformers Hugging Face library [81] for its demonstrated superior performance over GPT-3 and ChatGPT in capturing semantic relevance between sentences [82].

##### RAG: LLM Generator

Once all the necessary data was retrieved, we instructed the LLM (Gemini-2.0-Flash) to generate responses using the retrieved information. This process was guided by personalized prompts (PI, PE, PX) and the user need (i.e., informational, peer experiential expertise, and emotional responses) as described in Table 3. We iteratively refined the prompts via prompt engineering to improve comprehensiveness, accuracy, relevancy, transparency, and response coherence. The PI-2 prompt instructs the LLM to provide a detailed response followed by the PI-3 prompt to check the consistency of factual queries. For peer experiential expertise queries, we used the personalized experience (PX) prompt that instructs the LLM to use the retrieved documents and generate a third person narrative with explicit statements of partner conversations and direct quotes, when possible, to emphasize transparency.

For providing human-like emotional support, we used personalized emotion prompt-1 (PE-1) to first identify the feelings based on the 6 Willcox’s feeling categories and to generate a suitable response reflecting SSBC psychosocial constructs (i.e., sympathy, understanding, empathy, relief of blame, validation, compliment, or reassurance) for the identified feeling. Identifying user feelings allows responding in suitable emotional expressions that are typically associated with human responses. We also found that general LLM responses to emotional queries lacked deeper understanding of underlying emotions and human-like emotional behaviors, consistent with findings in previous literature [83]. The RAG LLM was instructed to use the style and tone from the retrieved emotional documents, which comprise peer to peer emotional support. When no relevant documents were retrieved for an emotional query, we used the PE-2 to generate a suitable response reflecting SSBC psychosocial constructs for the identified user feeling, without using the style and tone from the retrieved documents. Fact responses were validated by the fact validator. Then, the Cohesive Response (CR) prompt combines all subquery responses, instructing the LLM to smoothen the information flow into a single cohesive response. Finally, the LLM provides a combined response.

##### RAG: Fact Validator

Responses providing factual information were validated with the documents from the facts database, a step towards eliminating LLM hallucination. Our fact validation algorithm consists of two steps: fact identification and fact validation. To reduce overreliance on LLMs, we employed a deterministic technique to identify semantically relevant facts from the retrieved documents (i.e., the same top 5 documents retrieved from the Information Retriever process). This is done to address a known issue: that LLMs often provide imperfect fact identification when they break down complex information [89,90]. To identify the most relevant facts for validation, we (1) segmented the generated response and retrieved documents using the SAT tool and then (2) extracted the most relevant fact supporting document segment (Step 1) based on SBERT and cosine similarity (i.e., highest similarity score). This creates pairs of generated response segments that correspond to their fact supporting document segments. The LLM was instructed to use the response-document pairs and the FV-1 prompt (Step 2) to identify any response segments that are not supported by the document segments identified with cosine similarity. Providing the subset of relevant fact segments to the LLM narrows its focus, improving validation accuracy [1]. Then, using any generated response segments with inaccurate facts and the FV-2 prompt, the LLM was instructed to extract supporting facts from the retrieved documents (Step 3).

To evaluate the performance of the fact validator, we manually verified the performance for 1033 fact response segments from 100 generated responses for randomly sampled query responses that the LLM generated. The fact validator achieved a 97.39% validation accuracy based on 1006 sample items. The accuracy was calculated with the percentage of fact response segments that were fully supported by the facts database, as determined by the research team. Only 3% (n=27) of the generated response segments were not able to be validated with the facts database. These included response segments providing general information (n=12, e.g., *“Other side effects [of the medication] include changes in the immune system, dizziness, diarrhea, nausea, vomiting, headache, rash, and gas.”*) and personalized information (n = 8, e.g., *“Given the recent unprotected anal sex with someone who recently learned they had syphilis, your friend should get tested for STIs as soon as possible.”*). We also found false negatives (n=7), where the LLM extracted facts that could not support the generated response segment in Step 3. For example, when validating a response segment (e.g., *“Truvada alone is not sufficient for post-exposure prophylaxis (PEP).”*) with an accurate and semantically retrieved document segment, the LLM extracted facts that could support the generated response segment (e.g., *“It is worth noting that Truvada must be used correctly (without skipping doses) whether it is used in combination with other ARVs in the control of HIV, or for PrEP.”*). Based on these data, we opted for a three-step validation process, because the semantic retrieval (Step 1) identified over 82.10% (n=848) of accurate response segments, whereas the LLM identified (Step 2 and Step 3) 15.30% (n=158) of accurate response segments. It is also important to note that we also found that LLM-based validation was less consistent compared to the deterministic techniques. For instance, when we processed the same sets of 100 samples 10 times, the LLM changed its final validation responses (Step 2 and 3) in 44% of samples. It is important to note that the 44% does not indicate that earlier responses were inaccurate. Rather, the non-deterministic nature of LLMs (as probabilistic models) relying solely on LLM can hinder the validation process. This highlights the need for mixed validation approaches, combining deterministic techniques and LLMs to eliminate LLM hallucinations.

#### Internal Evaluation of the Chatbot Responses and Prototype Modification

The RAG chatbot was evaluated to assess the functional requirements following the iterative evaluation design process [84], with a focus on (1) accurately identifying user informational and emotional needs, (2) providing relevant and accurate information, and (3) generating personalized and emotionally suited responses. Developers conducted 10 rounds of internal evaluation of chatbot responses using randomly selected and paraphrased user queries from the experience database. Paraphrased user inputs from the experience database were used to create two test queries for each support need (i.e., informational, peer experiential expertise, and emotional). We selected (1) one simple and direct query and (2) one complex and context-dependent query. Responses were iteratively reviewed by at least 2 computational scientists and at least 1 HIV expert. Generated responses were evaluated based on the following 10 criteria: clarity [85,86], accuracy [82,85,87], actionability [87], relevancy [82,85], information detail [86], tailored information [82,85], comprehensiveness (questions answered fully) [86], language suitability [85], tone [86], and empathy [82]. The responses were also manually reviewed to verify that the chatbot only drew information from its database, ensuring the generated content aligns with retrieved documents.

These reviews were followed by chatbot modification to address issues identified by the reviewers. The most frequently occurring issues were: (1) low retrieval sensitivity and precision for context dependent queries (accuracy and comprehensiveness); (2) contradicting information from user discussions (accuracy), (3) response generation using inherent knowledge (accuracy), (4) irrelevant response context (relevancy), (5) fictional narratives (accuracy and transparency), (6) incomprehensive responses (comprehensiveness), and (7) noisy text (response coherence). Initially, LLM occasionally generated inaccurate information based on the user experience database. Thus, we refined the prompts to improve response quality. After we finalized the RAG database architecture, we followed LLM prompt engineering techniques [45,88]: we iteratively refined the response generation prompts (i.e., PI, PX, PE, FV, and CR) with a focus on improving response comprehensiveness, accuracy, relevancy, transparency, and coherence. First, we created level-1 prompts that were simple and direct instructions for generating a suitable response to a user query [88]. Then we refined the prompts to be more specific and structured to enhance information comprehensiveness, accuracy, relevancy, transparency, and response coherence.

All prompts were developed and refined sequentially and checked for overlapping effects. The process started with information subclassification (IC) and context handling prompts (CN-1 and CN-2). Then, we implemented personalized information prompts, followed by topic identification (PI-1), personalized response generation (PI-2), and verifying relevancy (PI-3). Then, the personalized experience prompt (PX) and personalized emotion prompts (PE-1, PE-2) were developed. Finally, we developed and refined the prompt for response cohesion (CR). The final prompts are shown in Table 3.

### Prompt Engineering for Information Comprehensiveness (Issues addressed: low retrieval sensitivity and precision, incomprehensive responses)

To improve information comprehensiveness, we refined the PI-2 prompt by instructing the LLM to segment the user query and provide a detailed response to all segments using the retrieved documents. In contrast, our level-1 PI-2 prompt (Appendix 1) simply instructed the LLM to provide a response to a user query using the retrieved documents; this generated response that were not comprehensive (i.e., the question was not answered fully).

### Prompt Engineering for Response Accuracy (Issues addressed: low retrieval sensitivity and precision, contradicting information, response generation using inherent knowledge, fictional narratives)

We refined our level-1 IC, CN-1, PI-2, PE-1, FV-1, FV-2, and PX prompts (Appendix 1) to improve the accuracy of generated responses. We refined the zero-shot IC prompt to a few-shot IC prompt to improve the accuracy of sub-classifying informational needs. The level-1 CN-1 prompt was iteratively paraphrased to verify whether additional context is needed for queries.

To enhance the accuracy of generated information, we refined the PI-2, FV-1, FV-2, and PX prompts to emphasize (by using ‘double asterisk’ ** in the LLM prompts) the use of only the retrieved documents for generating a response. Using the level-1 PI-2 prompt, the LLM still relied on inherent knowledge in responding to factual queries, even when instructed to use the retrieved factual documents to generate a response. The level-1 FV-1 and FV-2 prompts failed to identify facts that supported generated response segments. This was primarily due to a lack of clarity in defining factual consistency. This led to false positives, in which the LLM identified facts that only partially support the response segments. To prevent generating fictional real-world experiences, we refined the level-1PX prompt, which generated fictional self-narratives of HIV/PrEP experiences. This instructed the LLM to clearly indicate other people’s experiences or opinions as a third person narrative using the documents retrieved from the experience database.

To avoid inaccurate but anecdotal information in the experience database, we refined the PE-1 prompt to instruct the LLM to avoid using facts retrieved from the experience database to provide factual support. Although the level-1 PE-1 prompt included an instruction to use the retrieved documents only to reflect style and emotional tone, the LLM still generated factual support using the experience database. We found that the experience database included some inaccurate opinions. For example, one user posted *“…The risk, even if he is undetectable, isn’t zero, but it’s pretty low…”*, which contradicts the recognized medical information [110].

### Prompt Engineering for Information Relevancy (Issue addressed: irrelevant response context)

To strengthen information relevance, we developed the PI-3 prompt (See Table 3. for the final refined prompt) that verified the consistency of generated responses with the user query. The level-1 PI-2 prompt generated responses that lacked contextual awareness specifically regarding locations or populations. Refining the PI-2 prompt to have the additional task of verifying response consistency proved ineffective. This was a consistent theme in prompt engineering. To reduce the complexity of prompting, we developed a separate PI-3 prompt to verify consistency after generating a response using PI-2 prompt.

### Prompt Engineering for Information Transparency (Issue addressed: fictional narratives)

We refined the PX prompt by instructing LLM to include direct quotes of the user’s personal experiences when available and requiring the LLM to explicitly state that the response information was based on other people’s experiences (e.g., *“… This response is based on other people’s experience.”*).

### Prompt Engineering for Response Coherence (Issue addressed: noisy text)

To improve readability, comprehension, and user experience, we refined the PI-1, PI-2, PE-1, and PE-2 prompts (See Table 3. for the final refined prompts) to remove noisy responses, specifically unnecessary special characters (e.g., * in *“**Cost of PrEP in the US:**…”* and [ in *“[PrEP provided … infection.]”*) and logical reasoning, which included chatbot provided explanations for prompt execution (e.g., *“Okay, here are the revised responses, aiming for smoother flow and a more natural conversational tone:…”*) in responses.

#### Ongoing Development

##### Dataset Extension

To further improve the quality and diversity of data, we will expand the data sources to include two additional discussion boards from MedHelp and POZ community (HIV positive) to broaden the knowledge base. MedHelp and POZ databases consist of PrEP-related conversational dialogues among community members. We have collected these databases and will process the conversation dialogs to integrate in our RAG chatbot. We will develop prompts specifically tailored for varying formats and structures [91], including conversational dialogs, question-answer pairs, and dialogs with quotes. We will update the chatbot architecture to maintain historical dialogs to reflect real-world multi-turn conversations and include denial capabilities to set clear expectations for users and ensure safety compliance. We will also improve the fact validator’s accuracy to reduce the impact of LLM’s inherent inconsistencies when extracting facts to support generated responses. The final verification will focus on the accuracy of the responses rather than on the fact validator’s performance using the retrieved documents.

##### Chatbot Responses Quality and Formal Evaluation

To assess chatbot responses quality, an iterative formal evaluation will be conducted with 5-8 HIV experts. Eligible participants will have professional experience as an HIV counselor, researcher, health practitioner (e.g., HIV care providers), pharmacist, and/or social and community health worker. After providing online consent, participants will complete an online demographic questionnaire followed by the chatbot response evaluation questionnaire. The demographic questionnaire consists of 6 de-identified questions asking participants about their age, gender, ethnicity, race, region, professional title, years of experience as an HIV expert, and frequency of interaction with patients. The evaluation questionnaire will use a 5-point Likert scale to assess 10 evaluation criteria (Table 4) for 6 pairs of user queries and chatbot responses. Participants will have the opportunity to provide some additional information and human sample responses to help improve chatbot responses. Participants will receive a $25 Amazon gift card on completion of the study. IRB approval for the study is currently under review by [Blank for the review].

**Table 4.** Evaluation criteria and descriptions provided in chatbot response evaluation questionnaire.

| Attribute | Criterion | Description |
| --- | --- | --- |
| Information Quality | Information Accuracy [82,85,87] | Is the information provided factually correct and free from misinformation? |
|  | Information Clarity [85,86] | Is the information presented in a clear, concise, and easy-to-understand manner? |
|  | Information Action-ability [87] | Does the response provide practical, actionable advice that the user can implement? |
|  | Information Detail [86] | Does the response provide sufficient detail? |
| Response Relevance and Appropriateness | Relevance [82,85] | Does the response directly address the user's question and concerns? |
|  | Tailored Information [82,85] | Is the information provided tailored to the specific needs and circumstances of the user? |
|  | Comprehensiveness - Question Answered Fully [86] | Does the response fully answer the question without leaving significant gaps or requiring further clarification? |
| Language and Tone | Suitable Tone [86] | Is the tone of the response appropriate for the context and the user's overall emotional state? |
|  | Respectful, Empathetic, and Considerate Manner [82] | Does the response demonstrate empathy and understanding for the user's situation? Is the response respectful and non-judgmental? |
|  | Suitable Language [85] | Is the language free from jargon or technical terms that may be unfamiliar to the user? |

## Results

We have developed a RAG chatbot prototype as of Jan 2025 and have iteratively refined the chatbot design and system based on feedback from internal evaluation. We observed that prompt engineering plays a significant role in harnessing LLM capabilities to generate desired results. We report key findings from our iterative prompt engineering, offering suggestions for effective prompting in chatbot applications. Although there is no standard approach for refining prompts [94], starting with simple and direct instructions (Appendix 1) helped in understanding LLM comprehension and reasoning ability. For complex and longer queries, we found that prompting the LLM to decompose the query into segments helps in improving the response comprehensiveness (e.g., PI-2 prompt from Table 3). Such prompt decomposition was also effective in responding to subtle user emotions, by first identifying the emotions from a list of emotion categories and guiding the LLM in social-emotional reciprocation for personalizing emotional responses (i.e., PE-1 and PE-2 prompts).

An important finding in LLM prompt engineering was the limitation of using multiple instructions in a single prompt. We found that LLM often indicates low prompt adherence when a prompt has multiple instructions with logical reasoning to generate a desired response. Multiple, logic-based instructions can force frequent context switching, complicating LLM interpretation and response. Another challenge was the LLM’s sensitivity to linguistic variability in prompts [95]. We found that paraphrasing prompts by modifying prompt structure, using synonyms, and expanding prompt context, is ineffective in decision making tasks with high variability (e.g., identifying need of additional context). We found that condensed prompts with content words summarizing the main concept or idea, can reduce ambiguity in LLM decisions.

Although a few-shot prompting approach can be effective in text classification [45], we found that few-shot prompting did not consistently perform well across all classification tasks. Higher level classification tasks, such as classifying support needs, require diverse perspectives for inferencing ambiguous context and broad scope. Fine-tuning LLMs on annotated datasets outperformed few-shot prompting. Few-shot prompting is effective in simple sub-classification tasks with clear and distinct criteria for narrow scope [96].

For fact validation, deterministic techniques, including semantic relevance and text similarity metrics (e.g., ROUGE, BLEU) often failed due to their limited ability to capture negations [104] and numerical accuracy [108]. We found that LLMs generally struggle to validate statements that are generalized or personalized that are not directly present in the knowledge base. When required facts are scattered across documents, such generalized or personalized statements need implicit validation and cross referencing for accurate verification of information [103].

Results and analysis of expert opinions will be published in a subsequent manuscript.

## Discussion

We describe the development of a RAG chatbot for providing personalized support to fulfill informational, peer experiential expertise, and emotional needs of PrEP candidates to improve PrEP uptake. We hypothesize that the chatbot is capable of a). identifying support needs and personalizing responses (tailored information and clarity), b). providing complete, relevant and accurate HIV and PrEP information (comprehensiveness, relevancy, accuracy), c). providing peer experiential expertise support (relevancy, actionability), and d). responding with human-like emotional support (empathy). We expect our RAG chatbot can address limitations in current healthcare chatbots in several ways.

Our RAG chatbot can provide peer experiential expertise and human-like emotional support; this addresses gaps in current healthcare chatbots [121]. Experiential expertise support has a persuasive influence and is critical in empowering user decision making in health behaviors as it transfers lived experiences and action strategies to others in similar situations [55,60,97]. We ensured all the experiential responses explicitly stated that the information provided in the response was based on other people’s experiences, and restricted responses to emotional and peer experiential data from the experience database. Human-like emotional support using a widely validated Willcox’s Feeling Wheel as a framework [63] is a step closer to providing effective emotional support by identifying underlying user emotions. Such understanding is a critical element in improving HIV/PrEP behaviors [80].

Leveraging social media data helped address complex real-world user queries. RAG techniques enhance chatbot responses, reducing LLM hallucinations [44] and improving diversity of response content [53]. As opposed to fine-tuned LLMs, RAG chatbots can extract information from diverse sources to provide contextually intelligent [98], and comprehensive responses [53], and they are flexible to LLM updates. Using diverse sources of information in RAG can help integrate medical knowledge, peers’ experiences, as well as leverage social data to personalize and enhance chatbot provided health support.

LLM performance largely depends on the clarity and specificity of the instructions provided to perform a task [46,47]. Effective prompt engineering will guide LLM to generate the desired responses by iteratively refining the prompts based on response evaluation. The effectiveness of prompting strategies (e.g. tone, instruction format) varies with the complexity of the query and desired response. Using personalized and detailed prompts, the RAG chatbot can be instructed to focus on specific segments of user queries, clarify contextual ambiguity and leverage relevant documents to generate accurate and comprehensive responses compared to general LLM and traditional rule-based chatbots.

LLMs face challenges in logical reconstruction of fragmented information, in inferring nuanced statements [103], and in inherent inconsistencies (as probabilistic models) in generated output [109]. These challenges can hinder validation using LLM. To enhance the validation process, we leveraged the strength of both LLMs and traditional methods by minimizing dependence on LLMs for fact identification. We used this approach combined with LLM reasoning in cases where deterministic techniques fail. Although our approach improves validation performance, it is important to note that validating highly contextualized information is still on-going.

### Implications

RAG chatbots are increasingly used to provide various support for healthcare utilization and decision making [51,99–102]. We report the first published efforts to develop a RAG chatbot for providing personalized information, peer experiential expertise, and emotional support to PrEP candidates. Our chatbot improves PrEP support by leveraging verified information, real-world experiences, and emotional considerations. It can also interpret complex context-dependent queries and through personalized informational, peer experiential expertise, and human-like emotional support, we believe this approach can help reduce misconceptions about PrEP and HIV treatment, build user confidence, and promote PrEP uptake.

### Limitations

Although recent advances in hallucination removal have improved LLM reliability, these advances do not entirely eliminate the need for validating response accuracy, especially in healthcare applications [90]. Due to linguistic complexity and nuances in generated responses, LLMs struggle to consistently and accurately validate factual information using only RAGs [105]. Additional methodological improvements are needed to validate broad facts outside of the database and context-dependent information.

### Future work

We will analyze the ratings from experts for chatbot response quality across the 10 evaluation criteria and identify chatbot’s weak points via descriptive statistics (mean, standard deviation). To measure the consistency of ratings provided by the experts, we will calculate the intraclass correlation coefficient (ICC) [92] for the ratings of each criterion. We expect to have enough human sample responses after at least 3 rounds of evaluations. We will then compare the chatbot-generated responses to expert responses by measuring BLEU [82] and ROUGE-L [93] scores. These scores computationally measure relevance and linguistic similarity, and are scalable for evaluating larger datasets. In addition, human emotions are complex and are influenced by individual perception; this makes generating text for personalized emotional support a highly challenging endeavor. To ensure user safety and build trust and integrity, we will continue to refine our chatbot through a larger scale user study for gathering feedback on chatbot responses.

## Conclusion

This study details the implementation of a RAG chatbot designed to provide personalized informational, peer experiential expertise, and emotional support to PrEP candidates. By offering personalized information, peer experiences, essential information for making informed decisions in health, and human-like emotional support, our chatbot has the potential to reduce misconceptions and promote PrEP uptake. Expert and real-world feedback will validate and improve the chatbot’s potential; after this, a trial to demonstrate the efficacy of the tool to increase PrEP uptake and retention will be proposed.

## Data Availability

All data produced in the present study are available upon reasonable request to the authors

## Glossary

- Large Language Models – Artificial intelligence (AI) systems that are trained on large amounts of text data and capable of understanding and generating human-like responses [85].
- Natural Language Processing – A computational technique used to understand, interpret, and generate human language [101]
- Prompts: Instructions provided to an LLM to perform a specific task [45].
- Prompt Engineering – A technique used in developing and refining prompts to improve performance of LLMs in generating desired responses [45].
- Few-shot Prompting – A prompting technique used in LLMs where a few examples are provided in the prompts to guide the LLM in executing a specific task [88].
- Supervised Fine-tuning – A machine learning technique used to train a pre-trained LLM on a small dataset to adapt to domain specific tasks [113].
- Retrieval Augmented Generation – An AI technique that leverages an external knowledge base to improve the accuracy and relevancy of LLM generated responses [53].
- Hallucination or AI Hallucination– A phenomenon that refers to the generation of responses that seem plausible but are factually incorrect [43].
- Embeddings – Numerical representations of text data (words, phrases, or documents) that capture semantics and relationship between text elements for computational text processing [77].

## Appendix 1: Level 1 Prompt Engineering

**Table 1.**
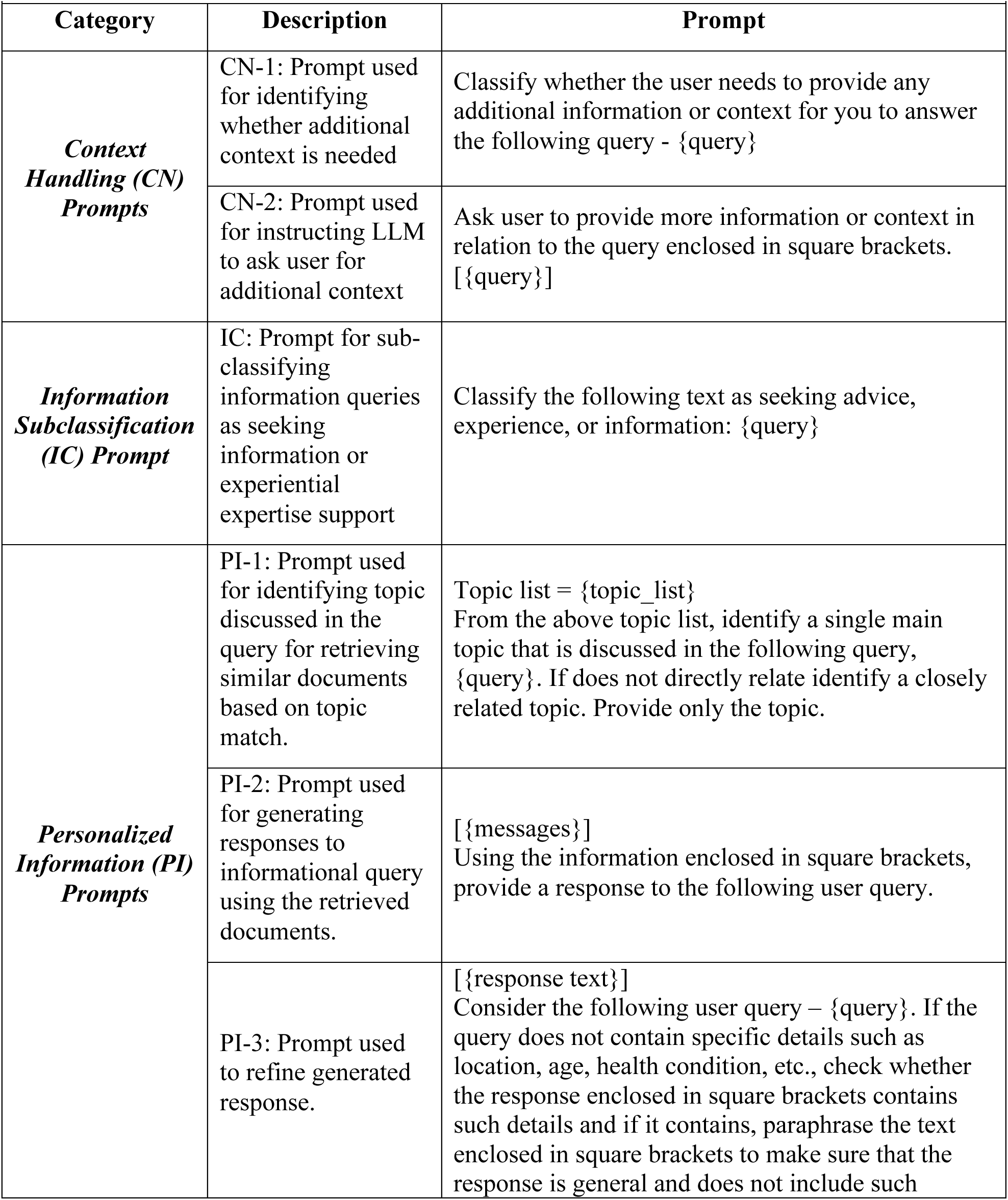
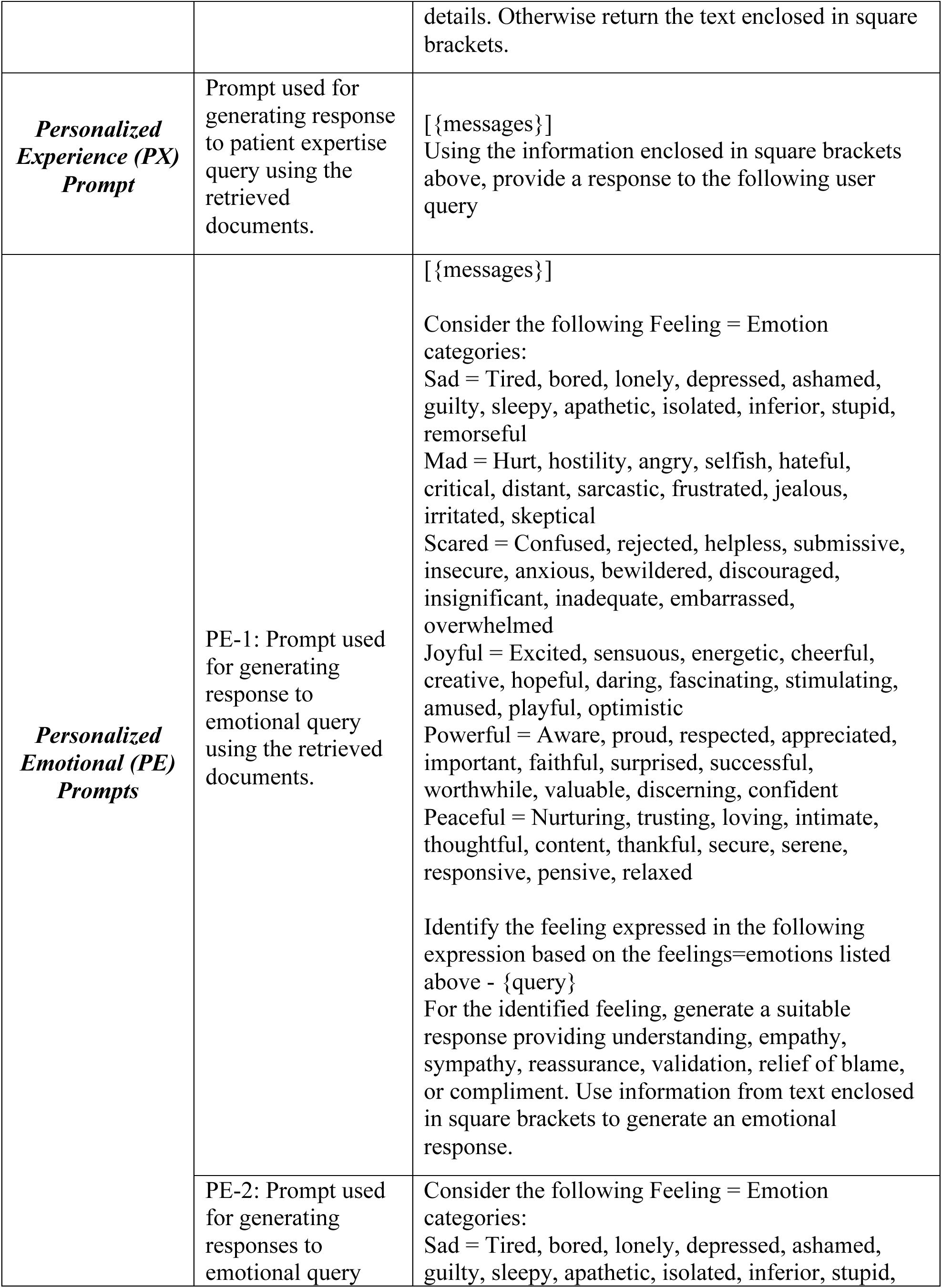
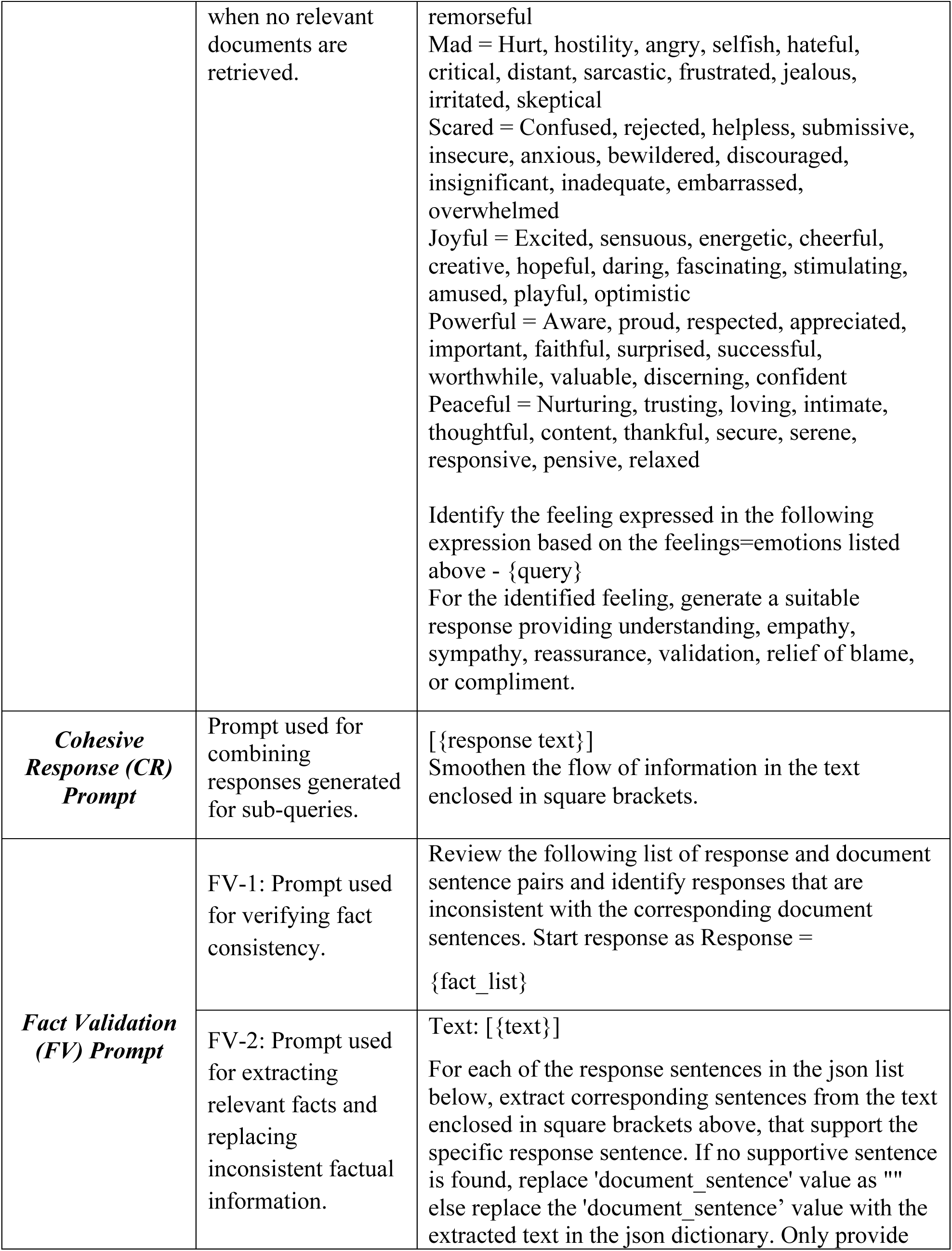
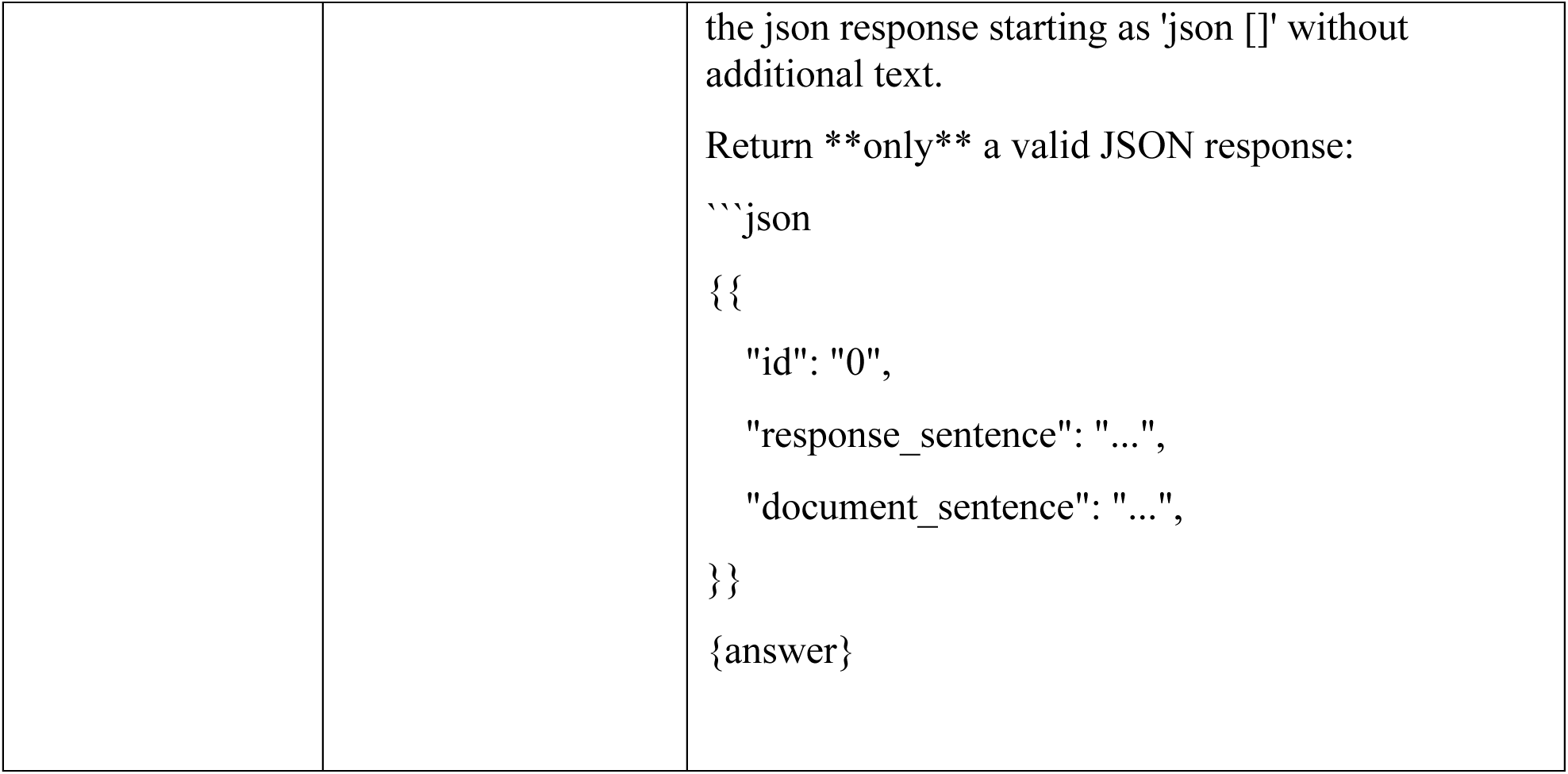
Initial prompts used for generating LLM responses to provide personalized PrEP support. These prompts were iteratively refined by developing several intermediate prompts to improve the quality of generated responses.

| Category | Description | Prompt |
| --- | --- | --- |
| **Context Handling (CN) Prompts** | CN-1: Prompt used for identifying whether additional context is needed | Classify whether the user needs to provide any additional information or context for you to answer the following query - {query} |
|  | CN-2: Prompt used for instructing LLM to ask user for additional context | Ask user to provide more information or context in relation to the query enclosed in square brackets. [{query}] |
| **Information Subclassification (IC) Prompt** | IC: Prompt for subclassifying information queries as seeking information or experiential expertise support | Classify the following text as seeking advice, experience, or information: {query} |
| **Personalized Information (PI) Prompts** | PI-1: Prompt used for identifying topic discussed in the query for retrieving similar documents based on topic match. | Topic list = {topic_list}<br>From the above topic list, identify a single main topic that is discussed in the following query, {query}. If does not directly relate identify a closely related topic. Provide only the topic. |
|  | PI-2: Prompt used for generating responses to informational query using the retrieved documents. | [{messages}]<br>Using the information enclosed in square brackets, provide a response to the following user query. |
|  | PI-3: Prompt used to refine generated response. | [{response text}]<br>Consider the following user query – {query}. If the query does not contain specific details such as location, age, health condition, etc., check whether the response enclosed in square brackets contains such details and if it contains, paraphrase the text enclosed in square brackets to make sure that the response is general and does not include such |
|  |  | details. Otherwise return the text enclosed in square brackets. |
| ***Personalized Experience (PX) Prompt*** | Prompt used for generating response to patient expertise query using the retrieved documents. | [ {messages} ]<br>Using the information enclosed in square brackets above, provide a response to the following user query |
| ***Personalized Emotional (PE) Prompts*** | PE-1: Prompt used for generating response to emotional query using the retrieved documents. | [ {messages} ]<br>Consider the following Feeling = Emotion categories:<br>Sad = Tired, bored, lonely, depressed, ashamed, guilty, sleepy, apathetic, isolated, inferior, stupid, remorseful<br>Mad = Hurt, hostility, angry, selfish, hateful, critical, distant, sarcastic, frustrated, jealous, irritated, skeptical<br>Scared = Confused, rejected, helpless, submissive, insecure, anxious, bewildered, discouraged, insignificant, inadequate, embarrassed, overwhelmed<br>Joyful = Excited, sensuous, energetic, cheerful, creative, hopeful, daring, fascinating, stimulating, amused, playful, optimistic<br>Powerful = Aware, proud, respected, appreciated, important, faithful, surprised, successful, worthwhile, valuable, discerning, confident<br>Peaceful = Nurturing, trusting, loving, intimate, thoughtful, content, thankful, secure, serene, responsive, pensive, relaxed<br>Identify the feeling expressed in the following expression based on the feelings=emotions listed above - {query}<br>For the identified feeling, generate a suitable response providing understanding, empathy, sympathy, reassurance, validation, relief of blame, or compliment. Use information from text enclosed in square brackets to generate an emotional response. |
|  | PE-2: Prompt used for generating responses to emotional query | Consider the following Feeling = Emotion categories:<br>Sad = Tired, bored, lonely, depressed, ashamed, guilty, sleepy, apathetic, isolated, inferior, stupid, |
|  | when no relevant documents are retrieved. | remorseful<br>Mad = Hurt, hostility, angry, selfish, hateful, critical, distant, sarcastic, frustrated, jealous, irritated, skeptical<br>Scared = Confused, rejected, helpless, submissive, insecure, anxious, bewildered, discouraged, insignificant, inadequate, embarrassed, overwhelmed<br>Joyful = Excited, sensuous, energetic, cheerful, creative, hopeful, daring, fascinating, stimulating, amused, playful, optimistic<br>Powerful = Aware, proud, respected, appreciated, important, faithful, surprised, successful, worthwhile, valuable, discerning, confident<br>Peaceful = Nurturing, trusting, loving, intimate, thoughtful, content, thankful, secure, serene, responsive, pensive, relaxed<br>Identify the feeling expressed in the following expression based on the feelings=emotions listed above - {query}<br>For the identified feeling, generate a suitable response providing understanding, empathy, sympathy, reassurance, validation, relief of blame, or compliment. |
| ***Cohesive Response (CR) Prompt*** | Prompt used for combining responses generated for sub-queries. | [ {response text} ]<br>Smoothen the flow of information in the text enclosed in square brackets. |
| ***Fact Validation (FV) Prompt*** | FV-1: Prompt used for verifying fact consistency. | Review the following list of response and document sentence pairs and identify responses that are inconsistent with the corresponding document sentences. Start response as Response = {fact_list} |
|  | FV-2: Prompt used for extracting relevant facts and replacing inconsistent factual information. | Text: [ {text} ]<br>For each of the response sentences in the json list below, extract corresponding sentences from the text enclosed in square brackets above, that support the specific response sentence. If no supportive sentence is found, replace 'document_sentence' value as "" else replace the 'document_sentence' value with the extracted text in the json dictionary. Only provide |
|  |  | the json response starting as 'json []' without additional text.<br>Return **\*\*only\*\*** a valid JSON response:<br>````json { "id": "0", "response_sentence": "...", "document_sentence": "...", } {answer}` |

